# Combining mpox vaccination and behavioural changes to control possible future mpox resurgence among men who have sex with men: a mathematical modelling study

**DOI:** 10.1101/2025.01.06.25320043

**Authors:** Maria Xiridou, Daphne Amanda van Wees, Philippe Adam, Fuminari Miura, Eline Op de Coul, Maarten Reitsema, John de Wit, Birgit van Benthem, Jacco Wallinga

**Author notes:** Corresponding author: Maria Xiridou, Centre for Infectious Diseases Control, Department of Epidemiology and Surveillance, National Institute of Public Health and the Environment, PO Box 1, 3720 BA Bilthoven, The Netherlands.

## Abstract

**Introduction:** The 2022 outbreak of mpox subclade IIb in the Netherlands affected primarily men who have sex with men (MSM). Despite the sharp decline in cases, there are concerns about future mpox outbreaks. We investigated the impact of mpox introductions, accounting for vaccination, behavioural changes, and introductions of subclades with different characteristics.

**Methods:** We developed a compartmental model for mpox among MSM. We distinguished three levels of sexual activity: low, medium, and high. The group with high activity was the 5% of the population with the highest numbers of sexual partners; the group with low activity was the 60% with the lowest numbers of partners. In the model, individuals were allowed to change sexual activity level. We examined several scenarios with mpox introductions.

**Results:** In January 2024, 54% of MSM with high sexual activity level had immunity; this declined to 39% a year later, due to changes in sexual activity. Introduction of 5 cases in May 2025 resulted in 759 cases in the first four months without vaccination after 2023, but the numbers of cases were 34% or 48% lower with 3,000 vaccinations in August-October 2024 or February-April 2025, respectively. With earlier initiation or greater magnitude of behavioural adaptations, the number of mpox cases was 17-51% smaller. Introduction of a subclade with 10% higher transmission probability than subclade IIb resulted in 77% more cases.

**Conclusions:** Due to changes in sexual activity, the fraction immune in the group with high sexual activity will decline, leading to greater possibilities for future mpox outbreaks. The number of cases can be reduced with preventive vaccination and behavioural adaptations. Recurring vaccination campaigns should prioritise individuals with high sexual activity level, ensuring those entering high-activity groups are reached. Campaigns promoting timely behavioural changes remain crucial.

**KEY MESSAGES:** *What is already known on this topic:* The numbers of mpox cases among men who have sex with men (MSM) in the Netherlands were low in 2023-2024, but there have been many introductions of new cases infected outside the Netherlands. Therefore, there are concerns about future mpox outbreaks, but it is uncertain whether new mpox vaccination campaigns are needed.

*What this study adds:* Due to behavioural changes, not-immune MSM enter the group with high sexual activity, thus reducing the fraction immune within the group and increasing the possibility of future mpox outbreaks. Preventive mpox vaccination and behavioural adaptations can impede mpox spread. However, outbreak sizes will increase as the time between vaccination roll-out and new introductions is extended.

*How this study might affect research, practice or policy:* Social marketing and interventions promoting timely behavioural changes and vaccination are essential. Recurring vaccination campaigns are necessary for mpox prevention, prioritising individuals with high sexual activity level and ensuring those entering high-activity groups are reached. The findings also emphasize that research is needed to understand determinants of behaviour and monitor behavioural changes over time.

## INTRODUCTION

In 2022-2023, there was a global mpox outbreak caused by subclade IIb, resulting in 93,030 mpox cases registered in 117 WHO Member States across all six WHO regions.^1^ Before this outbreak, there had been mpox outbreaks in the Democratic Republic of Congo and other African countries, mainly caused by mpox subclade Ia.^1–3^ Since 2023, there has been an increase in mpox cases and deaths in the Democratic Republic of Congo, due to subclade Ia and, most recently, also due to the novel subclade Ib.^1–4^ In 2024, travel-associated clade I cases were reported in several countries in Europe, North America, and Asia.^5–8^

The 2022 mpox outbreak in the Netherlands affected primarily men who have sex with men (MSM). The earliest date of symptom onset among confirmed mpox IIb cases was 27 April 2022.^9^ On 25 July 2022, the mpox preventive vaccination campaign started, targeting MSM and transgender persons most at risk of exposure to mpox.^11^ The 2022 mpox outbreak in the Netherlands showed a similar pattern as in various other European countries, but was characterized by a relatively high case rate per capita and possibly an earlier decline. The number of reported cases started declining before the start of vaccination, as the Dutch health authorities used a proactive communication strategy targeting MSM and sex venues early and effectively. ^9^ Additionally, the mandatory reporting of mpox cases in the Netherlands contributed to the completeness of mpox records, possibly explaining the high number of cases relative to the size of the MSM population.^10^ This provides reliable data to investigate the mpox situation in 2022-2023 and possible future resurgence.^9–11^ Furthermore, eligibility criteria for mpox vaccination were more restrictive than in other countries.^12^ The criteria in July 2022 were defined based on knowledge at that time about risk factors for mpox. The size of the target group was estimated at around 40,000 individuals.^10^ Approximately 18,000 individuals were vaccinated in the first round of preventive mpox vaccination (July 2022 – January 2023) and 1,500 in the second round (May-December 2023).^9–11^

The role of vaccination in future outbreaks depends on the level of immunity among the group of MSM with sexual activity that exposes them to possibilities of infection. However, defining this group is not straightforward, as evident by the changes in eligibility criteria for preventive mpox vaccination from 2022 to 2023. Preventive efforts are further complicated because individuals with high sexual activity levels who were infected or vaccinated in 2022-2023, may have lower sexual activity levels in the future, due to changes in behaviour. On the other hand, MSM with a low sexual activity level in 2022-2023 were probably neither infected nor vaccinated and, hence, they are not immune for mpox. If, due to changes in behaviour, these individuals have later higher sexual activity levels, the percentage immune in the highly-active group will decline. A study using data from the Amsterdam Cohort Studies investigating changes in the risk for HIV infection, found that MSM at low risk for HIV had a 0.11 probability of being at higher risk after 6 months,^13^ indicating that changes in the level of risk of an individual may occur over time.

Using a mathematical model for mpox among MSM, we investigated the impact of introductions of new mpox cases. We divided the MSM population into three subgroups with different levels of sexual activity, determined by the number of sexual partners. In the model, we allowed transitions of individuals from one activity group to another, due to changes in sexual behaviour. We examined the impact of vaccination and behavioural adaptations on the spread of mpox. The analyses were carried out with the characteristics of mpox subclade IIb, that was responsible for the 2022-2023 global outbreak. Due to the recent upsurge of subclade Ib,^14,15^ we also investigated the impact of introductions of cases infected with a (sub)clade with different transmissibility and severity than those of IIb.

## MATERIALS AND METHODS

### Transmission model

We developed a deterministic compartmental model for mpox among MSM (Figure S1). The model parameters are defined in Table 1 and Tables S1-S4 in the Supplement. We accounted for transmission via sexual or intimate contacts. In the model, MSM were divided into three groups, based on their level of sexual activity: low, medium, and high (Tables S1-S3 in Supplement). The division into the three activity groups was based on the number of sexual partners of MSM, obtained from data from the national registration of Sexual Health Centre (SHC) consultations.^11^ We allowed for transitions between the three activity groups, due to changes in sexual behaviour. For instance, MSM with a high sexual activity level may transition to the group with medium or low activity levels, if their number of sexual partners is reduced. The probabilities of transitions between these sexual activity groups were calculated with a methodology developed earlier^13,16^ (for details, see the Supplement and Tables S1-S3). To reflect uncertainty in parameter values, we explored parameter ranges and the model was calibrated to the daily number of mpox cases,^9^ according to the date of symptom onset, using a Bayesian approach (Figure S2 and methods in the Supplement). To examine the impact of transitions between sexual activity levels, we simulated the mpox outbreak of 2022-2023 under different parameter settings and repeated the calculations without transitions between the three sexual activity groups, such that MSM had the same sexual activity level throughout 2022-2023.

### Patient and public involvement

Patients or the public were not involved in the design, conduct, reporting, or dissemination plans for this study.

**Table 1.**
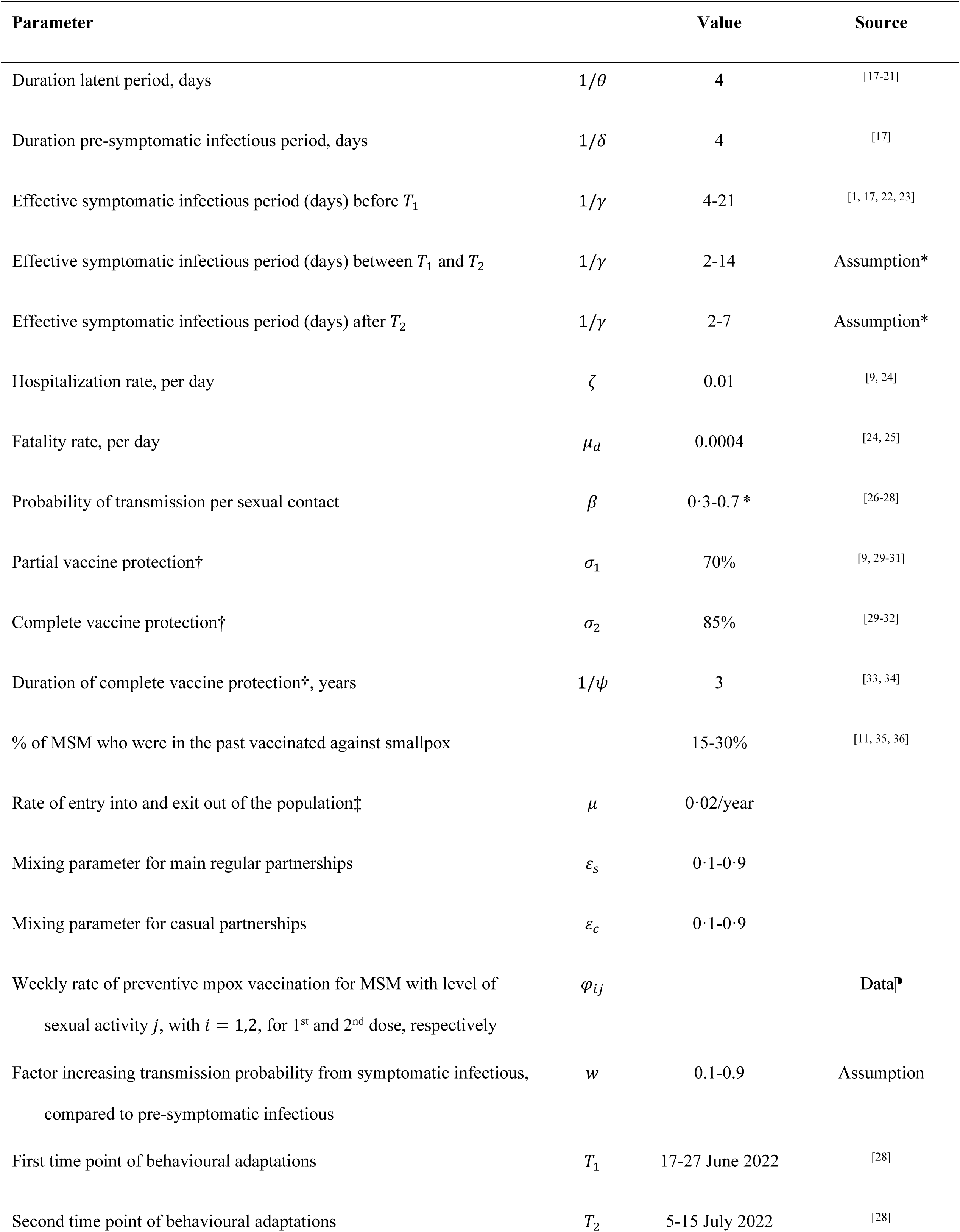
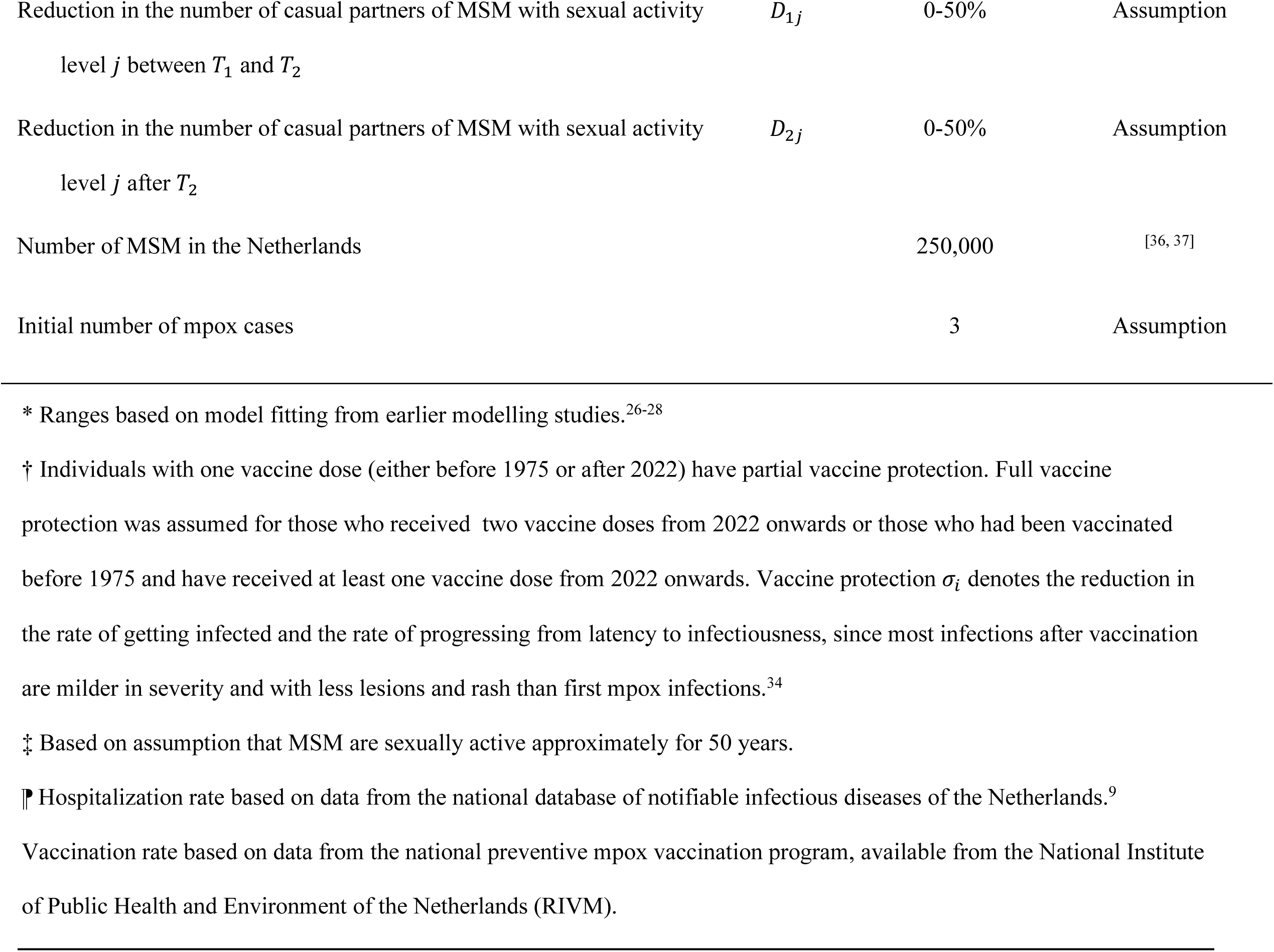
Model parameters. Ranges of values are given for parameters included in the uncertainty analysis.

### Mpox and vaccination

In the model, we distinguished four stages of mpox infection: exposed (not infectious), pre-symptomatic infectious, symptomatic infectious, and recovered/removed. We assumed that when symptomatic infectious cases start refraining from physical contacts (due to symptoms, knowledge of mpox, and/or advice from health professionals) or are hospitalized, they cannot infect others, and they transition to the compartment “recovered/removed”. MSM with past infection are assumed to be immune for the duration of the model calculations (approximately 4 years). MSM who received two vaccine doses in 2022-2023, as well as MSM who received both one vaccine dose in 2022-2023 and smallpox vaccination in the past (via the national smallpox vaccination program) are referred to as fully vaccinated. MSM who had only the old smallpox vaccination or only one vaccine dose in 2022-2023 are referred to as partially vaccinated. There is evidence that vaccine protection wanes over time;^38, 39^ therefore, individuals in the compartments “fully vaccinated” transition to the respective compartments “partially vaccinated” after 3 years. For simplicity, we did not account for waning protection for those partially vaccinated. The numbers of MSM vaccinated in 2022-2023 in the model were obtained from data from the National Institute of Public Health and the Environment^10, 11^ (Figure S2 in the Supplement).

### Behavioural adaptations in 2022

The model calculations started on 27 April 2022, the earliest date of symptom onset among confirmed mpox cases in the Netherlands. Earlier modelling work has shown that the 2022 outbreak in the Netherlands was greatly influenced by behavioural adaptations,^28^ that could have resulted from increased awareness and knowledge about mpox, public health authorities’ recommendations, perceived risk or perceived severity of mpox.^28,40^ Therefore, we also included in this study the possibility of behavioural adaptations: (a) a reduction in the number of sexual partners (for instance, due to increased awareness and perceived risk for mpox) and (b) a reduction in the effective infectious period (for instance, due to authorities’ recommendations to refrain from sexual contacts when diagnosed or due to increased knowledge about mpox thus recognising symptoms earlier). These adaptations were modelled to occur at two time points: in June and in July 2022. The timing and level of the adaptations were determined via a fitting process. We assumed that by the end of 2022, these adaptations were reversed, and behaviour was as before May 2022, since the mpox outbreak had waned.

### Scenarios for 2024-2026

We examined four scenarios based on the introduction of 5 or 10 new mpox cases on 1 May or 1 November 2025. The four scenario analyses were repeated five times: (a) without vaccination after 2023; (b-c) with 3,000 vaccinations carried out in August-October 2024 or February-April 2025; (d-e) with 30,000 vaccinations carried out in August-October 2024 or February-April 2025. The numbers of vaccinations were chosen to correspond to a small (3,000) or a large (30,000) vaccination program, reflecting experts’ recommendations to the Dutch government about possible mpox outbreaks and mpox vaccination in 2024-2025. For these scenarios, we assumed that the timing and levels of behavioural adaptations would be as in 2022. However, the knowledge and experience gained during the 2022 mpox outbreak may influence behaviours in the future. Behavioural adaptations might occur more quickly or be more pronounced. On the other hand, mpox may no longer be perceived as a significant threat and fatigue in responding to authorities’ recommendations could lead to delayed or less substantial adaptations. Therefore, we examined scenarios with behavioural adaptations occurring (a) 14 days earlier or 14 days later than in 2022; (b) at the same time point as in 2022, but the reductions in number of partners and in the duration of the effective infectious period were 25% higher or 25% lower than in 2022; and (c) when the number of symptomatic infectious cases exceeds 10 or 20 cases. Finally, we also examined two scenarios with introductions of mpox cases infected with subclades having different characteristics than those of subclade IIb: (a) 10% higher transmission probability per sexual act or (b) 50% higher hospitalization rate.^14,41–43^ The scenarios for 2024-2026 are summarized in Tables S5-S6 in the Supplement.

## RESULTS

### MSM with immunity to mpox

From the model, we calculated the percentage of MSM with immunity, including those who have had mpox, or mpox vaccination, or smallpox vaccination. By the end of 2023, when preventive mpox vaccination was stopped, the percentage of MSM with immunity was calculated at 30%, 34%, and 54% among MSM with low, medium, and high sexual activity level, respectively (Figure 1). We repeated the model calculations without transitions between sexual activity levels (assuming that all MSM remained with the same sexual activity level throughout 2022-2023); in that case, the respective percentages were 27%, 33%, and 89%. The difference between the two percentages in the subgroup with high sexual activity level reflects MSM who had a high sexual activity level at the beginning of the outbreak (and were mostly immune), but were later “replaced” by MSM who had a low sexual activity level at the beginning of the outbreak (and were mostly not immune). Due to the transitions between sexual activity groups, the percentage with immunity among those with a high sexual activity level declines over time to 39% on 1 January 2025 and 34% on 1 November 2025 (Figure 1).

**Figure 1.**
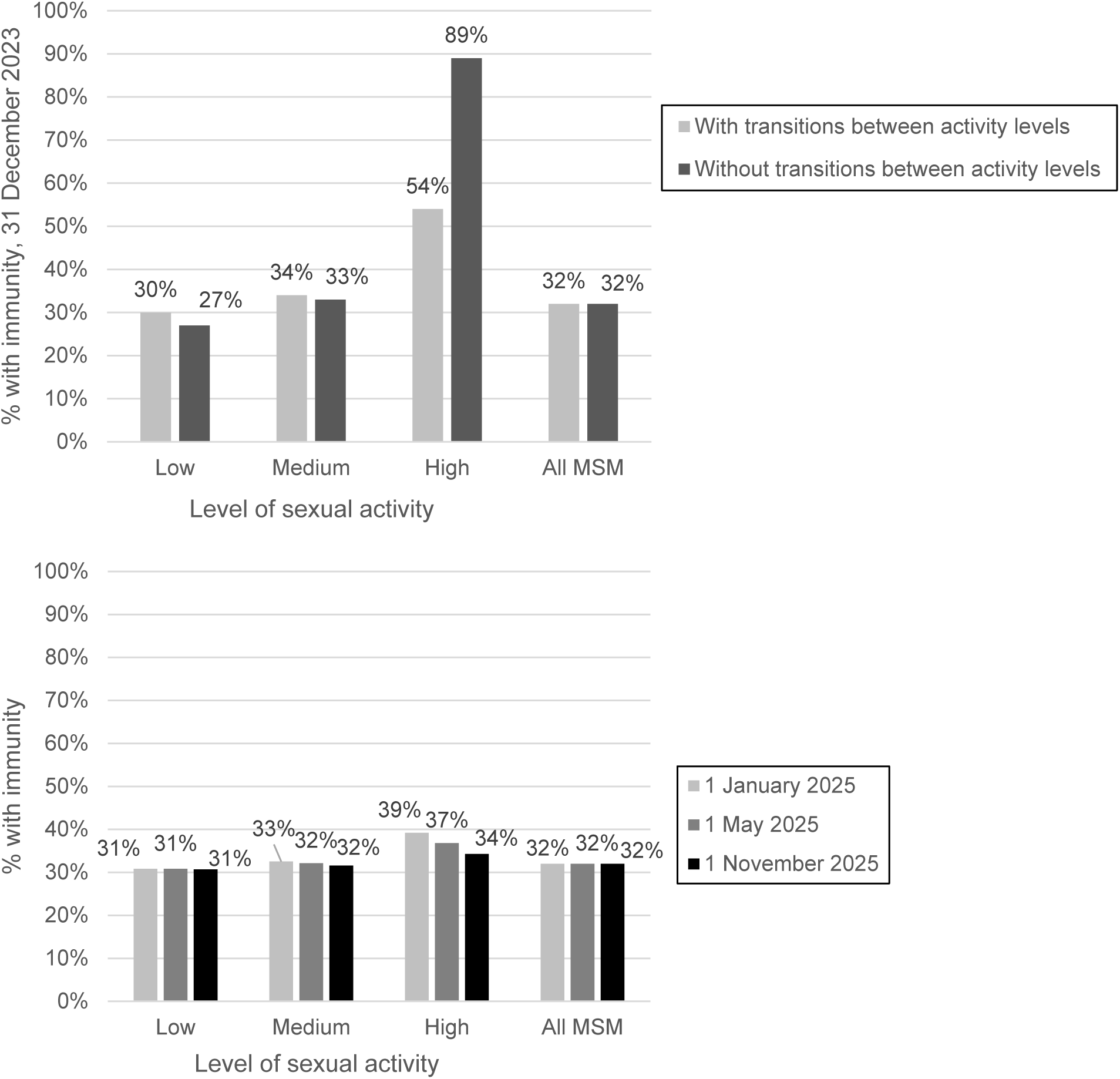
The percentage of MSM with immunity, as calculated from the model. Top panel: percentage immune on 31 December 2023; accounting for transitions of MSM between subgroups with different levels of sexual activity (grey) or without such transitions, where individuals remain with the same sexual activity level throughout 2022-2023 (black). Bottom panel: percentage immune on 1 January, 1 May, or 1 November 2025.

### New mpox introductions and the impact of vaccination

In the scenarios without additional vaccinations in 2024-2025, the introduction of 5 cases on 1 May 2025 resulted in an outbreak of 760 cases in the first 4 months and a peak of 16 daily cases (Figure 2 and Table S5). The introduction of the same number of cases half a year later (1 November 2025) resulted in more cases (860 in the first 4 months, with a peak of 18 daily cases), as the fraction with immunity among MSM with high sexual activity level declined over time (Figure 1). With 3,000 vaccinations, the number of cases was smaller, compared to the scenario without additional vaccinations. However, as the period between vaccination and introduction of new cases became longer, the number of cases increased, with a total of 502 cases in the first 4 months if vaccination was carried out in August-October 2024, compared to a total of 395 cases, if vaccination was carried out in February-April 2025. With 30,000 vaccinations, there were very few secondary cases, especially in the scenarios where index cases were introduced short after vaccination was carried out. Similar findings were obtained in the scenarios based on the introduction of 10 new mpox cases (Figure S3 in the Supplement), which resulted though in almost twice as many mpox cases, compared to the scenarios with 5 new introductions (Figures 2 and S3, Table S5).

**Figure 2.**
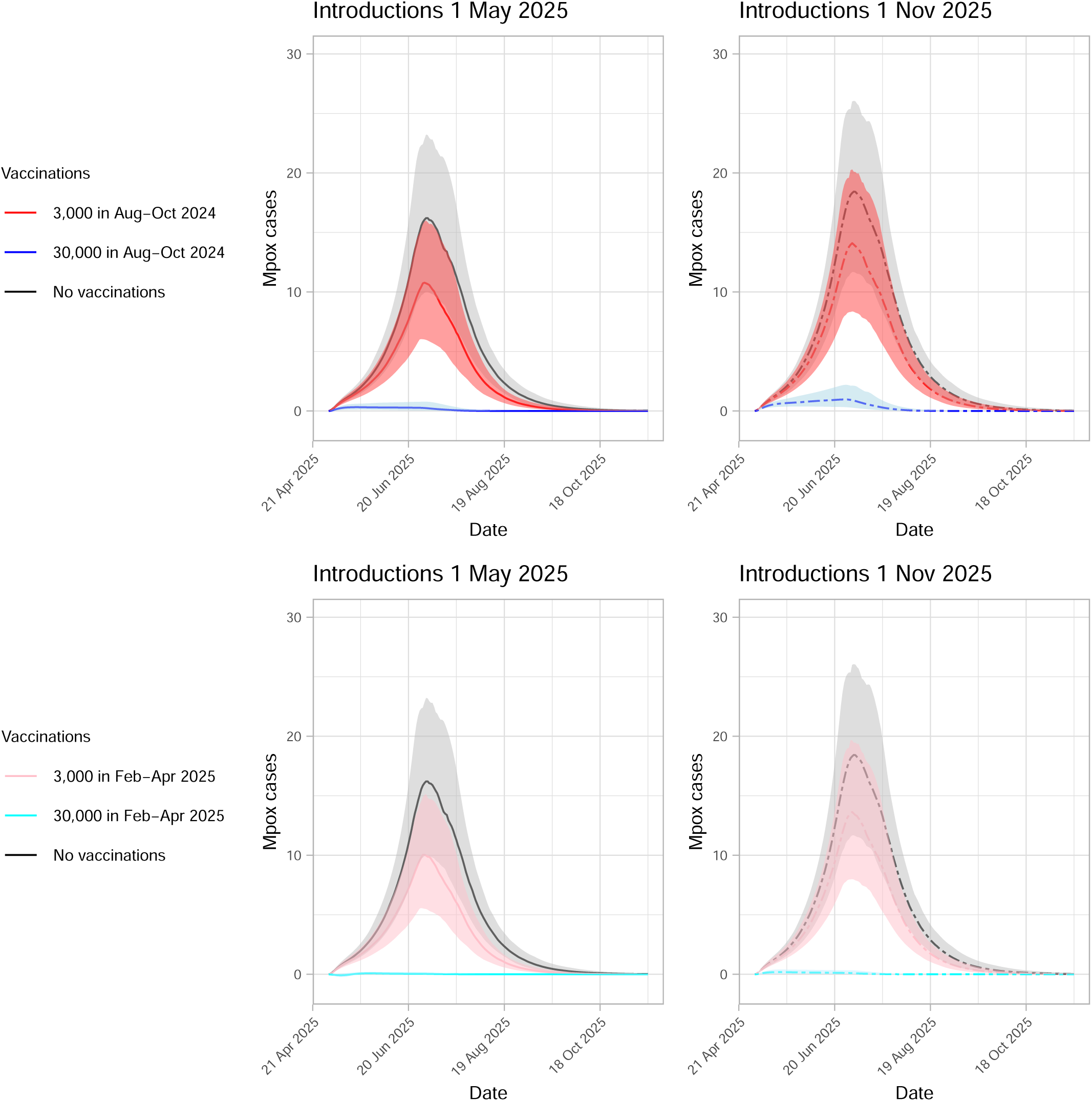
Mpox outbreaks after the introduction of 5 new (index) mpox cases on 1 May (left panels; solid lines) or 1 November 2025 (right panels; dashed lines). The lines show the medians and the shaded areas show the 95% credible intervals for the scenarios with the respective colours. Black lines and grey shaded areas: without vaccinations after 2023. Top panels: red lines and shaded areas: with 3,000 vaccinations carried out in August-October 2024; dark blue lines and shaded areas: with 30,000 vaccinations carried out in August-October 2024. Bottom panels: pink lines and shaded areas: with 3,000 vaccinations carried out in February-April 2025; light blue lines and shaded areas: with 30,000 vaccinations carried out in February-April 2025. The vaccinations in 2024 or 2025 were uniformly distributed over the 3-month period, resulting in 250 or 2,500 vaccinations per week, for the scenarios with total 3,000 or 30,000 vaccinations, respectively.

### Different behavioural adaptations, compared to the 2022 outbreak

In the scenarios where the behavioural adaptations started 14 days earlier or the level of reduction was higher, the number of mpox cases was lower, compared to the respective scenario with behavioural adaptations occurring as in 2022: for instance, 51% and 17% lower, respectively, with introductions on 1 May 2025 (Figure 3, Table S6). When the reductions started later or the level of reduction was smaller than in 2022, the number of mpox cases was higher: for instance, 82% or 32% higher, respectively, with introductions on 1 May 2025. With behavioural adaptations occurring when the number of symptomatic cases exceeded 10 or 20 cases (Figure S4 in Supplement), the number of mpox cases was considerably lower than with adaptations as in 2022: 92% or 84% lower, respectively.

**Figure 3.**
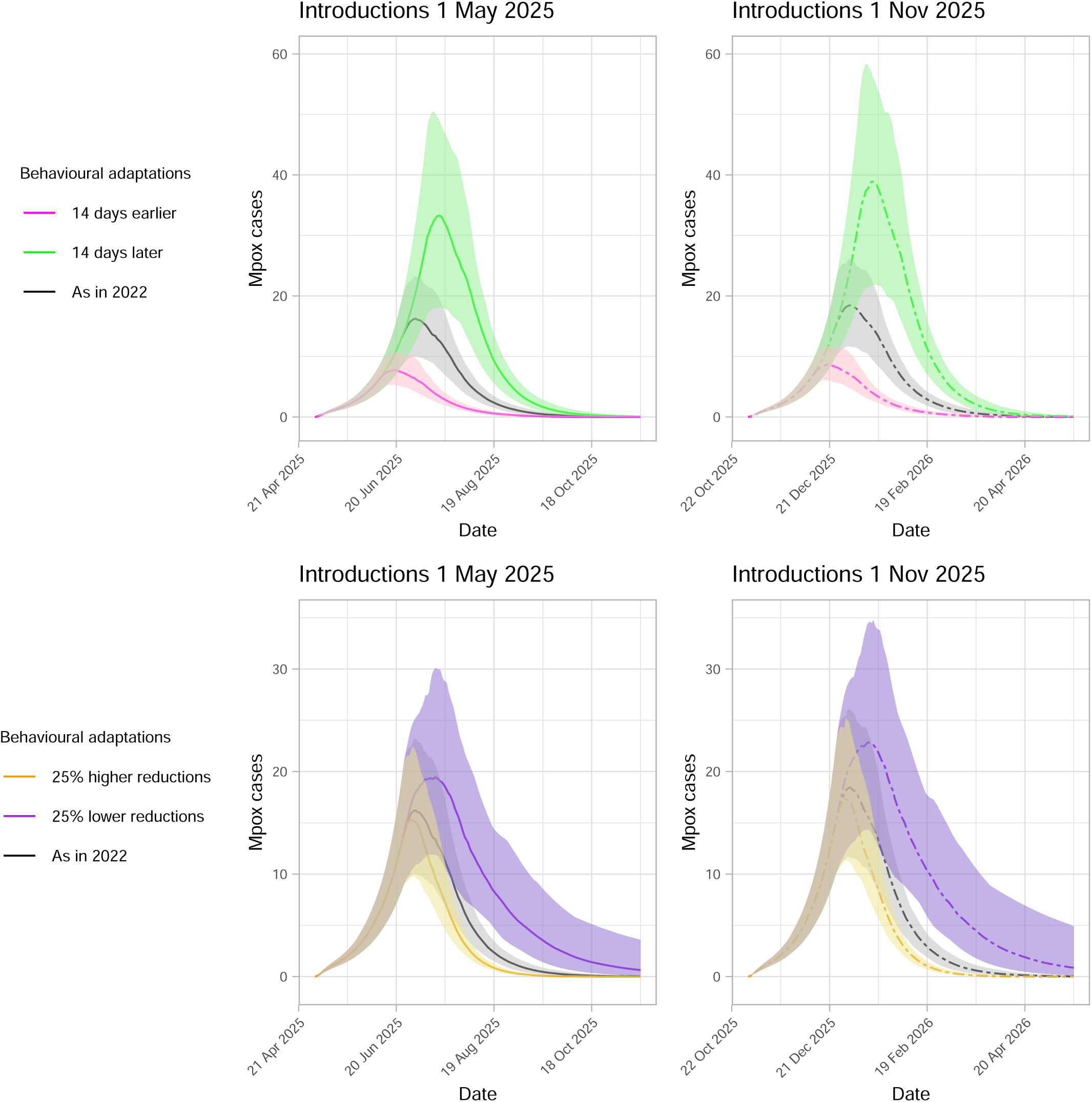
Mpox outbreaks after the introduction of 5 new (index) mpox cases on 1 May (left panels; solid lines) or 1 November 2025 (right panels; dashed lines) without vaccinations after 2023. The lines show the medians and the shaded areas show the 95% credible intervals for the scenarios with the respective colours. Black lines and grey shaded areas show the scenarios with behavioural adaptations as during the 2022 outbreak, starting approximately 50 days after the earliest day of symptom onset. Top panels: Behavioural adaptations started 14 days later (green lines and shaded areas) or 14 days earlier (purple lines and shaded areas), compared to the 2022 outbreak. Bottom panels: The reduction in the number of partners and the reduction in the effective infectious period were 25% lower (purple lines and shaded areas) or 25% higher (yellow lines and shaded areas), than those in 2022.

### Introduction of mpox cases infected with a different subclade

Introduction of 5 mpox cases on 1 May 2025 resulted in 82% more cases, when the newly introduced cases were infected with a subclade with 25% higher transmissibility than subclade IIb (Figure 4 and Table S6). Introduction of cases infected with a subclade with higher hospitalization rates resulted in 74% less cases, assuming that those hospitalized do not have contacts and hence do not infect others. Similar results were observed in scenarios based on the introduction of 10 or 20 mpox cases infected with a different (sub)clade (Figure S5 in Supplement).

**Figure 4.**
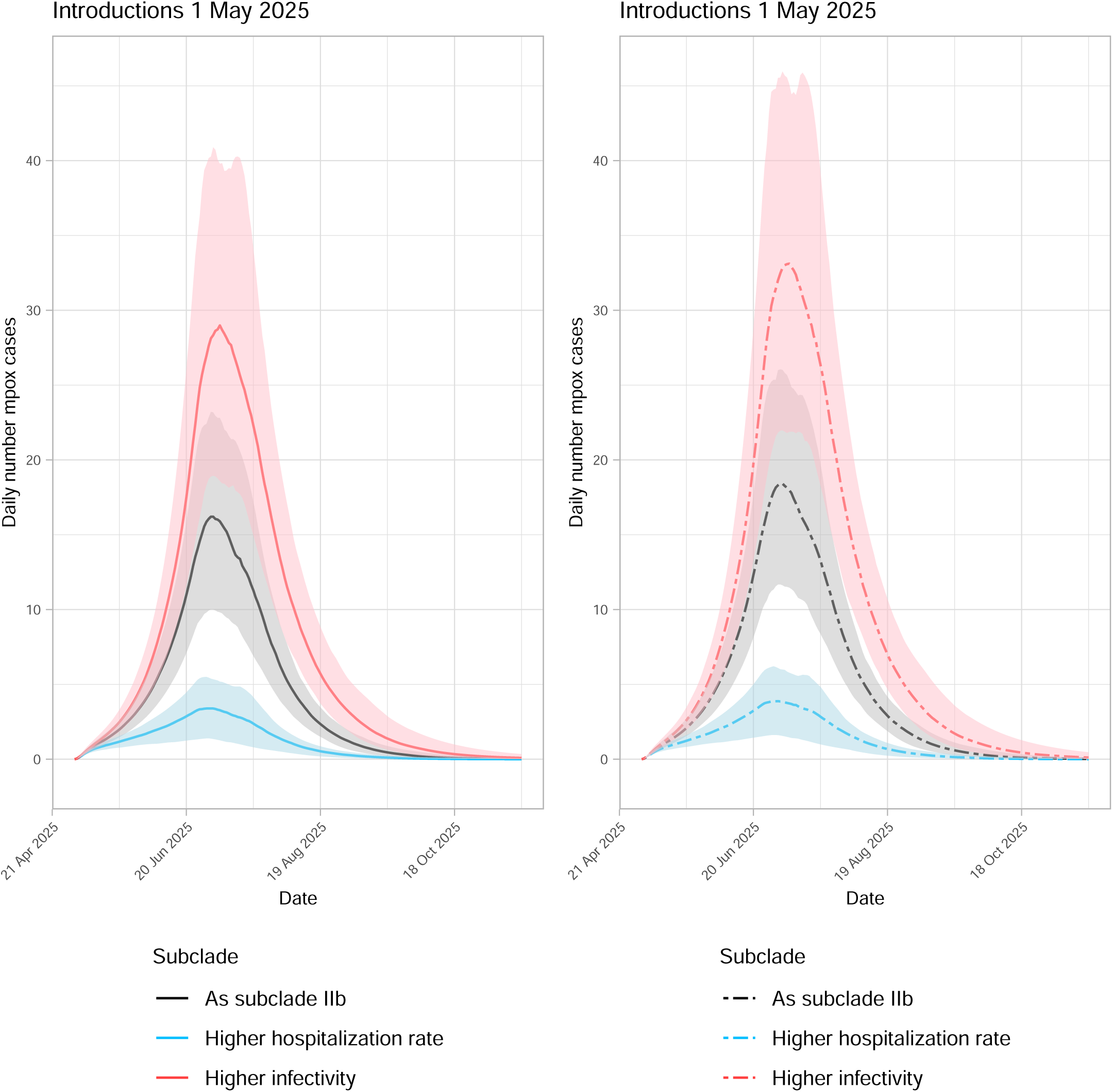
Mpox outbreaks after the introduction of 5 new mpox cases on 1 May (left panel; solid lines) or 1 November 2025 (right panel; dashed lines), without additional vaccinations after 2023. The lines show the medians and the shaded areas show the 95% credible intervals for the scenarios with the respective colours. The new cases were infected with a clade having: the same properties as clade IIb, responsible for the 2022 outbreaks (black lines and grey shaded areas); or with higher infectivity (red lines and pink shaded areas); or with higher hospitalization rate (blue lines and shaded areas).

## DISCUSSION

In this study, we estimated that when preventive mpox vaccination was stopped in the Netherlands by the end of 2023, more than half of MSM with a high sexual activity level had immunity via infection or vaccination.

However, this fraction may be declining over time as MSM with low sexual activity levels in 2022, who are likely still uninfected and unvaccinated, may have higher sexual activity levels in later years. In our analyses, future introductions of mpox cases resulted in a higher number of secondary cases than the same number of introductions at an earlier time point, due to the declining fraction of immune individuals among those with a high sexual activity level. In the scenarios including 3,000 vaccinations in 2024-2025, mpox spread was reduced compared to the scenarios without vaccinations after 2023, but with longer time intervals between roll-out of vaccination and introduction of new cases, the number of mpox cases in future outbreaks was higher. When we included a much higher number of 30,000 vaccinations in 2024-2025, new mpox introductions resulted in only limited transmission. Furthermore, we found that behavioural adaptations strongly influenced the size of future outbreaks: when we incorporated early initiation or great magnitude of behavioural adaptations, the number of secondary mpox cases was substantially smaller. In analyses accounting for introduction of new cases infected with a subclade that is more transmissible than subclade IIb, we found higher numbers of mpox cases, compared to the respective scenarios for subclade IIb. A subclade with higher hospitalization rate resulted in less mpox cases, assuming that those hospitalized did not infect others.

The findings of this study highlight the necessity to maintain a high level of immunity among MSM with high sexual activity levels. Therefore, preventive vaccination strategies should prioritise those with high sexual activity, ensuring that individuals newly entering the high-activity group are promptly vaccinated. Booster vaccinations of those vaccinated a few years earlier could also contribute to increasing the level of immunity. This underscores the need to provide opportunities to receive mpox vaccination again, either via a campaign or via structural availability of the vaccine. Interventions targeting those at risk for mpox should consider that individuals who are not immune to mpox may enter this group due to modifications in their behaviour, thus reducing the fraction immune in the group at risk. Recurring vaccination campaigns (for instance, repeated annually) of a few thousand MSM with high sexual activity may be of great value to control mpox spread. Our results stress the significance of timely behavioural adaptations and the importance of accurate information enabling individuals to adapt their behaviour in order to protect themselves and their partners, but also information that facilitates the decision to vaccinate. To this end, social marketing campaigns and other measures, aiming to help people make timely positive decisions and/or changes can play a crucial role.

Our findings regarding the importance of behavioural adaptations in response to a future outbreak are in agreement with our earlier modelling study showing that the 2022 outbreak in the Netherlands was shaped by behavioural adaptations.^28^ This was corroborated by modelling studies from other countries.^21,26,44,45^ Modelling studies of possible future mpox outbreaks have also shown that vaccination and behavioural interventions could considerably reduce the size of future outbreaks of subclade IIb^46^ or Ib.^47^

The transmission model used in this study is simple, yet it accounts for the most important factors that determine mpox dynamics: different stages of mpox infection, different levels of sexual activity, different vaccination statuses, and behavioural adaptations in response to an outbreak. A strength of our study is that we explicitly incorporated transitions of MSM between subgroups with different sexual activity levels. This feature allows us to project how these transitions affect the level of immunity in the subgroup with high sexual activity, and consequently, the size of future outbreaks and the impact of preventive vaccination. Furthermore, the model is informed by observed epidemiological and behavioural data on sexual behaviour, as well as the reported numbers of vaccinations and mpox cases in the Netherlands.

A limitation of this study is that the level of sexual activity was based only on the number of sexual partners. Other factors, such ashaving sex in sex venues or sex parties also determine the level of sexual activity and have been shown to be independent risk factors for mpox diagnosis.^48^ Furthermore, we did not account for differences in behaviour according to HIV-status or PrEP use. Also, we did not incorporate age-stratification in the model, due to lack of data to estimate model parameters according to age. However, due to age-assortative mixing, the level of immunity among very young MSM may have remained lower than among older MSM. Another limitation of the study is that the estimated transition rates of MSM between subgroups may be biased due to a possible overrepresentation of MSM with a higher number of sexual partners in the data we used. Data from the national registration of SHC consultations were used in this study, because longitudinal observation of sexual behaviour per person is required to estimate the transition rates, but was not available from other sources. Furthermore, we did not account for the underreporting of mpox infections or for infections remaining asymptomatic until recovery, due to lack of data to assess whether and to what extent this occurred in the Netherlands. Studies from other countries have reported small numbers of asymptomatic or undetected infections,^49–52^ and we expect that this simplification in our study does not considerably affect our findings qualitatively. Additionally, it should be considered that barriers to seeking mpox testing or vaccination, including stigma,^53,54^ could hinder the implementation of interventions as modelled in this study, thus reducing their potential benefits. Due to lack of data, we made several assumptions about uncertain parameters, such as the duration of immunity after infection or vaccination, the characteristics of different mpox subclades, or the level of awareness among MSM and how that could influence their behaviour. It would be beneficial to obtain data that enable a more accurate estimation of these characteristics. Finally, our study aimed to present credible intervals to reflect the range of parameter uncertainty and visualize plausible epidemic trajectories under different scenarios, rather than full predictive uncertainty, including observation noise. It is possible though that our approach may have underestimated uncertainty due to the model structure and distributional assumptions.

Our model can be expanded to include age-stratification and account for heterogeneity according to age, HIV status, or PrEP use. Future research could investigate the impact of these forms of heterogeneity on mpox transmission. Other directions for future work would be to examine mpox transmission also via heterosexual, household, or other contacts, as well as the influence of the structure of the contact network on the transmission dynamics of mpox. The impact and cost-effectiveness of vaccination and booster vaccination of those infected or vaccinated in the past could also be investigated in future work.

In conclusion, our study reveals that the possibility of an mpox outbreak will increase over time, despite the experienced infections and vaccinations thus far, as not-immune individuals enter the group with high sexual activity. Our findings highlight the importance of a combination of vaccination and behavioural adaptations to control future outbreaks. The severity of the outbreaks could be reduced with early behavioural adaptations of considerable magnitude. Conversely, the outbreak size may increase as the interval between vaccination campaigns and new mpox introductions is extended, since uninfected-and-unvaccinated individuals may enter the group with high sexual activity, thus reducing the fraction immune within this group. It is imperative to maintain a high level of immunity among those with high sexual activity, indicating the importance of recurring vaccination campaigns targeting this group. Social marketing and other interventions aiming to assist individuals make timely positive decisions about their behaviours are essential. Surveillance and preparedness for possible future outbreaks remain crucial.

## STATEMENTS

### Patient consent

Not applicable.

### Ethical approval

Not applicable.

### Data availability

Data with the numbers of daily mpox cases in the Netherlands are freely available on the website of the National Institute of Public Health and Environment of the Netherlands: https://www.rivm.nl/en/mpox/current-information-about-mpox. The weekly numbers of mpox vaccinations are available in the report “Sexually transmitted infections in the Netherlands in 2023” (page 152) of Kayaert L, et al. (RIVM report 2024-0038; Center for Infectious Diseases Control, National Institute of Public Health and the Environment (RIVM); Bilthoven, 2024. Available online: https://www.rivm.nl/bibliotheek/rapporten/2024-0038.pdf). Data from the PrEP pilot are not available for replication, because they are third-party data and are not freely available. They are data from the Dutch national registration of Sexual Health Centre consultations (SOAP). Pseudonymized individual participant data can be requested for scientific use with a methodologically sound proposal submitted to the SOAP registration committee for approval. Information can be requested from the authors. The code for the model analyses is available at https://github.com/rivm-syso/Mpox-Vac-RiskChanges.

## Acknowledgements

The authors would like to thank Manon Haverkate for the data on mpox vaccinations, Laura Kayaert for data on mpox cases, Danja Sarink for assistance with the data for the analyses on transitions between risk groups, Ilja van Bergen for suggestions about the impact of behaviour, Jantien Backer and Marten van Antwerpen for assistance with R code, Hester Korthals-Altes for assistance with modelling the mpox risk groups.

