## Supplementary File for "Combining mpox vaccination and behavioural changes to control possible future mpox resurgence among men who have sex with men: a mathematical modelling study"

### SUPPLEMENT

### The transmission model

We developed a deterministic compartmental model that describes monkeypox virus (MPXV) transmission among men who have sex with men (MSM), based on our earlier work [1]. The model structure is shown in Figure S1 in the main text. We accounted only for transmission via sexual or intimate contacts between regular and casual sex partners. In this paper, the term "regular partner" refers to a person with whom an individual engages in regular or ongoing sexual activity, typically (but not exclusively) within a committed relationship. Sex partners other than regular partners are referred to as casual partners in this study. The MSM population was divided into subgroups based on three categorizations:

- Level of sexual activity: low, medium, or high.
- Mpox status: uninfected, exposed (not infectious), infectious pre-symptomatic, infectious symptomatic, or recovered/removed (not infecting others).
- Vaccination status: unvaccinated, partially vaccinated, or fully vaccinated.

These categories are explained in detail in the following paragraphs.

### The course of MPXV infection

Mpox symptoms can begin a few days after exposure and typically last 2-4 weeks [2-4]. There is evidence that transmission can occur before recognizable symptoms appear [5-7]. The symptomatic infectious period ends when people recover from mpox ("recovered") or when they are hospitalized or start refraining from physical contacts ("removed" from the group that can infect others). Individuals with mpox may refrain from physical contacts due to symptoms, knowledge of mpox, and/or advise from health professionals [8-10]. This means that the actual infectious period (the period with virus shedding) is practically reduced to the effective infectious period (the period during which individuals are infectious and have contacts that enable transmission). Individuals enter the population as susceptible ($S_{ij}$) when they become sexually active. After infection, we distinguished four stages of mpox infection: exposed (but not infectious: $E_{ij}$), pre-symptomatic infectious ($I_{ij}$) [5-7], symptomatic infectious ($Y_{ij}$), and recovered or removed ($R_{ij}$). The subscripts $i$ denote the vaccination status and the subscripts $j$ denote the level of mpox risk; these are explained in the following paragraphs. The pre-symptomatic phase lasts $1/\delta$ days after becoming infectious. The effective symptomatic infectious period lasts $1/\gamma$ days after symptom onset. These are assumed to be the same for all subgroups.

**Subgroups of MSM with different levels of sexual activity**

The MSM population was divided into $G = 3$ subgroups with different levels of sexual activity. For brevity, we refer to these subgroups as sexual activity groups or activity groups. The division into the three activity groups was based on data of MSM participating at the PrEP pilot from July 2019 (start of the PrEP pilot) until December 2022 (data obtained from the Dutch national registration of Sexual Health Centre consultations (SOAP) [11]). The participants of the PrEP pilot visit the Sexual Health Centres every three months for PrEP consultations, according to the Dutch PrEP guidelines [12]. During these consultations, they provide information about their sexual behaviour in the previous 6 months. We used the number of sex partners in the preceding 6 months that was reported at each consultation as the main characteristic to determine the level of sexual activity. We considered the 5% with the highest numbers of sex partners among all consultations as the subgroup with high sexual activity and the 60% with the lowest numbers of sex partners as the subgroup with low sexual activity; the remaining 35% was defined as medium activity level. The characteristics of the three groups are shown in Tables S1-S2. Subsequently, we modelled transitions of MSM from one activity group to another as a Markov process and we obtained six-month transition probabilities between the three levels of sexual activity (Table S3). The procedure to define the activity groups and to calculate the transition probabilities between activity groups has been described in detail in earlier publications [13, 14]. The level of sexual activity is denoted with the second subscript of the classes $S_{ij}$, $E_{ij}$, $I_{ij}$, $Y_{ij}$, $R_{ij}$, with $j = 1$ denoting the low level, $j = 2$ denoting medium level, and $j = 3$ denoting high level of sexual activity, respectively.

**Vaccination status**

Smallpox vaccination was ended in the Netherlands in 1975. Therefore, when the mpox outbreak started in April 2022, a fraction of the MSM population was vaccinated via historical smallpox vaccination, that offered partial protection against mpox [15, 16]. In 2022-2023, MSM at risk for mpox received preventive mpox vaccination [17]. According to national guidelines, individuals who had received smallpox vaccination in the past would receive one vaccine dose for mpox, while unvaccinated individuals would receive two vaccine doses for mpox. In the model, vaccination status was distinguished as:

- Unvaccinated: those who have never had smallpox or mpox vaccination.

- Partially vaccinated: MSM who have received historical smallpox vaccination (before 1975) and no mpox vaccination; MSM who had not received smallpox vaccination and had only one mpox vaccine dose in 2022-2023.
- Fully vaccinated: MSM who had received smallpox vaccination and one mpox vaccine dose in 2022-2023; MSM who had two mpox vaccine doses in 2022-2023.

The vaccination status is denoted with the first subscript of the classes $S_{ij}$, $E_{ij}$, $I_{ij}$, $Y_{ij}$, $R_{ij}$, where $i = 0$ denotes unvaccinated, $i = 1$ is for partially vaccinated, and $i = 2$ for individuals fully vaccinated. We assumed that protection with two doses lasts $1/\psi$ years after the most recent vaccine dose. After that period, those fully vaccinated are partially vaccinated. Partially vaccinated individuals who receive a new vaccine dose are fully vaccinated for $1/\psi$ years. It takes two weeks from vaccine administration until the individual is actually protected and we accounted for no vaccine protection during these two weeks. To account for this two-week interval, the number of MSM vaccinated in week $t$ in the model was the number of MSM who received vaccination in week $t - 2$ in the national database of mpox vaccinations.

Vaccine protection is denoted with $\sigma_1$ for those partially vaccinated and $\sigma_2$ for those fully vaccinated. We assumed that vaccination protects against infection and against disease. This means that among individuals partially or fully vaccinated ($i = 1,2$, respectively): only a fraction $1 - \sigma_i$ of uninfected may get infected with MPXV and only a fraction $1 - \sigma_i$ of exposed may develop mpox. Vaccine protection is higher for those fully vaccinated than for those partially vaccinated ($\sigma_2 > \sigma_1$). Uninfected individuals may be vaccinated at a rate $\varphi_{ij}$, where $i$ denotes the vaccination status and $j$ denotes the level of sexual activity. All MSM entering the sexually active population are unvaccinated and uninfected.

**Model equations**

The model is described by the system of ordinary differential equations shown below. The following vectors of state variables are defined:

- $\boldsymbol{S}_i = \{S_{i1}, S_{i2}, S_{i3}\}$ are the numbers of susceptibles with vaccination status $i$ in sexual activity group 1,2,3.
- $\boldsymbol{E}_i = \{E_{i1}, E_{i2}, E_{i3}\}$ are the numbers of exposed with vaccination status $i$ in sexual activity group 1,2,3.
- $\boldsymbol{I}_i = \{I_{i1}, I_{i2}, I_{i3}\}$ are the numbers of pre-symptomatic infectious with vaccination status $i$ in sexual activity group 1,2,3.

- $\boldsymbol{Y}_i = \{Y_{i1}, Y_{i2}, Y_{i3}\}$ are the numbers of symptomatic infectious individuals with vaccination status $i$ in sexual activity group 1,2,3.
- $\boldsymbol{R}_i = \{R_{i1}, R_{i2}, R_{i3}\}$ are the numbers of recovered/removed individuals with vaccination status $i$ in sexual activity group 1,2,3.

We use bold letters for vectors and matrices; symbols not in bold denote numbers. Let $\boldsymbol{C}$ denote the 3x3 matrix with the transition probabilities between the three sexual activity groups shown in Table S3 and $\boldsymbol{J}$ a 3x1 vector with all elements equal to 1. We use the symbol · for matrix multiplication and the symbol ° for Hadamard products (element-wise products) between matrices or between vectors with the same dimensions.

Equations for unvaccinated individuals:

$$\frac{d\boldsymbol{S}_0}{dt} = -(\boldsymbol{\lambda} + \boldsymbol{\varphi}_0 + \mu)^\circ \boldsymbol{S}_0 + \mu \boldsymbol{N} - (\boldsymbol{C} \cdot \boldsymbol{J})^\circ \boldsymbol{S}_0 + \boldsymbol{S}_0 \cdot \boldsymbol{C}$$

$$\frac{d\boldsymbol{E}_0}{dt} = \boldsymbol{\lambda}^\circ \boldsymbol{S}_0 - (\theta + \mu)\boldsymbol{E}_0 - (\boldsymbol{C} \cdot \boldsymbol{J})^\circ \boldsymbol{E}_0 + \boldsymbol{E}_0 \cdot \boldsymbol{C}$$

$$\frac{d\boldsymbol{I}_0}{dt} = \theta \boldsymbol{E}_0 - (\delta + \mu)\boldsymbol{I}_0 - (\boldsymbol{C} \cdot \boldsymbol{J})^\circ \boldsymbol{I}_0 + \boldsymbol{I}_0 \cdot \boldsymbol{C}$$

$$\frac{d\boldsymbol{Y}_0}{dt} = \delta \boldsymbol{I}_0 - (\gamma + \zeta + \mu + \mu_d)\boldsymbol{Y}_0 - (\boldsymbol{C} \cdot \boldsymbol{J})^\circ \boldsymbol{Y}_0 + \boldsymbol{Y}_0 \cdot \boldsymbol{C}$$

$$\frac{d\boldsymbol{R}_0}{dt} = (\gamma + \zeta)\boldsymbol{Y}_0 - \mu \boldsymbol{R_0} - (\boldsymbol{C} \cdot \boldsymbol{J})^\circ \boldsymbol{R}_0 + \boldsymbol{R}_0 \cdot \boldsymbol{C}$$

Equations for partially vaccinated individuals:

$$\frac{d\boldsymbol{S}_1}{dt} = -[(1 - \sigma_1)\boldsymbol{\lambda} + \boldsymbol{\varphi}_1 + \mu]^\circ \boldsymbol{S}_1 + \boldsymbol{\varphi}_0{}^\circ \boldsymbol{S}_0 + \psi \boldsymbol{S}_2 - (\boldsymbol{C} \cdot \boldsymbol{J})^\circ \boldsymbol{S}_1 + \boldsymbol{S}_1 \cdot \boldsymbol{C}$$

$$\frac{d\boldsymbol{E}_1}{dt} = (1 - \sigma_1)\boldsymbol{\lambda}^\circ \boldsymbol{S}_1 - (\theta + \mu)\boldsymbol{E}_1 - (\boldsymbol{C} \cdot \boldsymbol{J})^\circ \boldsymbol{E}_1 + \boldsymbol{E}_1 \cdot \boldsymbol{C}$$

$$\frac{dI_1}{dt} = (1 - \sigma_1)\theta \boldsymbol{E}_1 - (\delta + \mu)\boldsymbol{I}_1 - (\boldsymbol{C} \cdot \boldsymbol{J})^\circ \boldsymbol{I}_1 + \boldsymbol{I}_1 \cdot \boldsymbol{C}$$

$$\frac{d\boldsymbol{Y}_1}{dt} = \delta \boldsymbol{I}_1 - (\gamma + \zeta + \mu + \mu_d)\boldsymbol{Y}_1 - (\boldsymbol{C} \cdot \boldsymbol{J})^\circ \boldsymbol{Y}_1 + \boldsymbol{Y}_1 \cdot \boldsymbol{C}$$

$$\frac{d\boldsymbol{R}_1}{dt} = (\gamma + \zeta)\boldsymbol{Y}_1 + \sigma_1\theta\boldsymbol{E}_1 - \mu\boldsymbol{R}_1 - (\boldsymbol{C}\cdot\boldsymbol{J})^\circ\boldsymbol{R}_1 + \boldsymbol{R}_1\cdot\boldsymbol{C}$$

Equations for fully vaccinated individuals:

$$\frac{d\boldsymbol{S}_2}{dt} = -[(1-\sigma_2)\boldsymbol{\lambda} + \psi + \mu]^\circ\boldsymbol{S}_2 + \boldsymbol{\varphi}_1{}^\circ\boldsymbol{S}_1 - (\boldsymbol{C}\cdot\boldsymbol{J})^\circ\boldsymbol{S}_2 + \boldsymbol{S}_2\cdot\boldsymbol{C}$$

$$\frac{d\boldsymbol{E}_2}{dt} = (1-\sigma_2)\boldsymbol{\lambda}^\circ\boldsymbol{S}_2 - (\theta + \mu)\boldsymbol{E}_2 - (\boldsymbol{C}\cdot\boldsymbol{J})^\circ\boldsymbol{E}_2 + \boldsymbol{E}_2\cdot\boldsymbol{C}$$

$$\frac{d\boldsymbol{I}_2}{dt} = (1-\sigma_2)\theta\boldsymbol{E}_2 - (\delta + \mu)\boldsymbol{I}_2 - (\boldsymbol{C}\cdot\boldsymbol{J})^\circ\boldsymbol{I}_2 + \boldsymbol{I}_2\cdot\boldsymbol{C}$$

$$\frac{d\boldsymbol{Y}_2}{dt} = \delta\boldsymbol{I}_2 - (\gamma + \zeta + \mu + \mu_d)\boldsymbol{Y}_2 - (\boldsymbol{C}\cdot\boldsymbol{J})^\circ\boldsymbol{Y}_2 + \boldsymbol{Y}_2\cdot\boldsymbol{C}$$

$$\frac{d\boldsymbol{R}_2}{dt} = (\gamma + \zeta)\boldsymbol{Y}_2 + \sigma_2\theta\boldsymbol{E}_2 - \mu\boldsymbol{R}_2 - (\boldsymbol{C}\cdot\boldsymbol{J})^\circ\boldsymbol{R}_2 + \boldsymbol{R}_2\cdot\boldsymbol{C}$$

The parameters and variables in these equations are explained in the following sections, in Table 1 in the main text, and Tables S1-S3.

**Transmission rate**

The rate at which MSM in sexual activity group $j$ get infected with MPXV is $\lambda_j = \lambda_{rj} + \lambda_{cj}$, where $\lambda_{rj}$ and $\lambda_{cj}$ denote the rates of getting infected by regular and casual partners, respectively:

$$\lambda_{rj} = q_j \sum_{i=0}^{2}\sum_{\hat{j}=1}^{3} m_{rj\hat{j}}[1-(1-\beta)^{u_{j\hat{j}}}]\frac{wI_{i\hat{j}}+Y_{i\hat{j}}}{N_{\hat{j}}} \quad \text{and} \quad \lambda_{cj} = \alpha_{cj}\beta\sum_{i=0}^{2}\sum_{\hat{j}=1}^{3} m_{cj\hat{j}}\frac{wI_{i\hat{j}}+Y_{i\hat{j}}}{N_{\hat{j}}}$$

In matrix form, $\boldsymbol{\lambda} = \boldsymbol{\lambda}_r + \boldsymbol{\lambda}_c$ with:

$$\boldsymbol{\lambda}_r = \boldsymbol{q}^\circ[(\boldsymbol{M_r}{}^\circ\boldsymbol{B})\cdot\boldsymbol{Z})] \text{ and } \boldsymbol{\lambda}_c = \beta\boldsymbol{\alpha}_c{}^\circ(\boldsymbol{M_c}\cdot\boldsymbol{Z})$$

where $\boldsymbol{Z} = [w(\boldsymbol{I}_0 + \boldsymbol{I}_1 + \boldsymbol{I}_2) + \boldsymbol{Y}_0 + \boldsymbol{Y}_1 + \boldsymbol{Y}_2]/\boldsymbol{N}$ (element-wise division) and $\boldsymbol{B}$ is a 3x3 matrix with the $\{j,k\}$-elements equal to $1-(1-\beta)^{u_{jk}}$. In these equations, the following notation is used:

- $\boldsymbol{q} = \{q_1, q_2, q_3\}$ are the fractions of MSM of sexual activity group 1,2,3 with a regular partner.
- $u_{j\hat{j}}$ is the frequency of sex contacts between regular sexual partners of sexual activity groups $j, \hat{j}$, calculated as $u_{j\hat{j}} = (u_j + u_{\hat{j}})/2$, from the contact frequency $u_j$ and $u_{\hat{j}}$ of groups $j$ and $\hat{j}$, respectively.

- $w$ is a factor reducing the transmission probability from a pre-symptomatic infectious individual compared to symptomatic infectious individuals.
- $\boldsymbol{\alpha_c} = \{\alpha_{c1}, \alpha_{c2}, \alpha_{c3}\}$ and $\alpha_{cj}$ is the number of casual sex contacts per day for men in sexual activity group $j$.
- $\boldsymbol{N} = \{N_1, N_2, N_3\}$ are the sizes of the three sexual activity groups and $N_T = \sum_{j=1}^{G} N_j$ is the total size of the MSM population.
- $\boldsymbol{M_r}$ and $\boldsymbol{M_c}$ are 3x3 matrices with elements $m_{rij}$ and $m_{cij}$, respectively, defining the level of mixing between sexual activity groups $i, j$, when forming regular and casual partnerships, as follows:

$$m_{rij} = \varepsilon_r \delta_{ij} + (1 - \varepsilon_r) \frac{\alpha_{rj} N_j}{\sum_{v=1}^{G} \alpha_{rv} N_v} \quad \text{and} \quad m_{cij} = \varepsilon_c \delta_{ij} + (1 - \varepsilon_c) \frac{\alpha_{cj} N_j}{\sum_{v=1}^{G} \alpha_{cv} N_v},$$

where $\delta_{ij}$ is the Kronecker delta (being equal to 1, if $i = j$; and equal to 0, otherwise) and the parameters $\varepsilon_r$, $\varepsilon_c$ determine the level of assortativeness in mixing of activity groups when forming regular and casual partnerships, respectively (if $\varepsilon_i = 1$, then mixing is assortative; if $\varepsilon_i = 0$, then mixing is proportionate). This implies that

$$\boldsymbol{M}_r = \varepsilon_r \boldsymbol{I_3} + \frac{\mathbf{1} - \varepsilon_r}{\boldsymbol{\alpha_r} \cdot \boldsymbol{N}} [(\boldsymbol{\alpha_r} \circ \boldsymbol{N}) \cdot \boldsymbol{J}]^T \quad \text{and} \quad \boldsymbol{M}_c = \varepsilon_c \boldsymbol{I_3} + \frac{\mathbf{1} - \varepsilon_c}{\boldsymbol{\alpha_c} \cdot \boldsymbol{N}} [(\boldsymbol{\alpha_c} \circ \boldsymbol{N}) \cdot \boldsymbol{J}]^T,$$

where $\boldsymbol{I_3}$ is the 3x3 identity matrix and $\boldsymbol{A}^T$ denotes the transpose of $\boldsymbol{A}$.

**Historical smallpox vaccination**

To estimate the percentage of MSM who had been vaccinated before 1975, via the historical smallpox vaccination program, we assumed that individuals born in 1974 or earlier were vaccinated, while those born in 1975 and thereafter were not vaccinated. The percentage of the MSM population born until 1974 was based on the following data, which were available in categories (for instance, 40-44 or 40-49 years old):

- Statistics Netherlands provides data for the male population of the Netherlands in 2022; this is the total population, not only MSM. According to this database, there were 5,715,970 men aged 15-65, of whom 32% was older than 50 and 42% was older than 45 [18].
- 25.7% of those tested positive for mpox and 25.4% of those tested negative for mpox in the Netherlands until 3 April 2023 was ≥45 years old [19].

- Among individuals visiting Sexual Health Centra in the Netherlands, 18.5% were ≥50 years old [20].
- In an online survey about mpox among MSM, 34.3% of respondents was ≥50 years old [21].
- Among MSM visiting the Sexual Health Centra of Rotterdam and Amsterdam, 13.6% and 12.3%, respectively, were ≥50 years old [22].

Based on these data, we assumed that approximately 25% of the MSM population in April 2022, had received smallpox vaccination in the past.

**Behavioural adaptations in response to the mpox outbreak in 2022**

In May-July 2022, there was considerable attention in news and social media about the outbreak and health authorities provided information and recommendations that could help individuals reduce their risk to get infected [1, 21, 23-25]. Therefore, it is possible that, in the first weeks of the outbreak, MSM adapted their behaviour in response to the outbreak, due to the increased awareness, authorities' recommendations, perceived risk and perceived severity of mpox [1, 24, 26]. A number of surveys in several countries found evidence of adaptations to sexual behaviour in response to the mpox outbreaks, such as reducing numbers of sexual partners and less frequently visiting sex venues [27-29]. In surveys carried out a few weeks after the start of the outbreaks, MSM reported high levels of knowledge, awareness, but also worries and concerns about mpox and the mpox outbreak [27-30]. Earlier modelling work in the Netherlands has pointed to the importance of behavioural adaptations in June and July 2022 in controlling the mpox outbreak [1]. Modelling studies in other countries also corroborate this finding [31-34].

Therefore, in the model we accounted for the possibility of behavioural adaptations in response to the mpox outbreak of 2022. We included: (a) a reduction in the number of casual partners and (b) a reduction in the effective symptomatic infectious period, due to earlier refraining from sexual contacts among those with symptoms. The level and the timing of the adaptations were obtained from the fitting process. The first time point of behavioural adaptations was $T_1$ and it was sampled from the uniform distribution in the range 17-27 June 2022. The second time point of behavioural adaptations was $T_2$ and was sampled from the uniform distribution in the range 5-15 July 2022. The reduction in the number of casual partners was $D_{1j}$ between $T_1$ and $T_2$, while the reduction was $D_{2j}$ after $T_2$, with $j = 1,2,3$ denoting MSM with low, medium, or high sexual activity, respectively. The six values $D_{ij}$ were sampled from the uniform distribution in the range 0-50%.

#### Model fitting and uncertainty analysis

We used data on the daily numbers of mpox cases from the national surveillance system for notifiable infectious diseases [15]. The numbers of confirmed mpox cases were ordered according to date of symptom onset.
The model was calibrated to the number of mpox cases using a Bayesian approach. To reflect uncertainty in model parameters, we explored wide ranges of values and defined uniform prior distributions for the uncertain parameters (Table 1 in main text, Table S1). Using Latin Hypercube Sampling [35], we sampled 100,000 combinations of parameter values. We started the model calculations with three infectious unvaccinated cases in the group of MSM with high risk for mpox. With each combination of parameter values, we computed the daily number of mpox cases and its Poisson likelihood until September 2022, because after that date there were only sporadic cases in the Netherlands [15, 36]. This provided the posterior distributions of the uncertain parameters (Table S4). The model results and the data are shown in Figure S2.

#### Data sources

The following data sources were used in this study.

*National surveillance system for notifiable diseases (OSIRIS):* Individuals seeking mpox testing or presenting with symptoms suggestive of mpox to general practitioners or health centra were referred and notified to the regional public health service for mpox diagnostics [15]. Notification of confirmed mpox cases from regional public health services to the National Institute of Public Health and the Environment was accompanied by a questionnaire with demographical, clinical, and epidemiological information of cases. The numbers of confirmed mpox cases according to date of symptom onset were extracted from this database. These numbers are continuously updated and are available online on https://www.rivm.nl/mpox-apenpokken.

*Dutch national registration of Sexual Health Centre consultations (SOAP):* In this database, surveillance data of all STI consultations from all 24 STI clinics in the Netherlands are accumulated [11]. Data of those participating in the PrEP pilot are also registered in this database. The PrEP pilot started in July 2019. During the three-monthly PrEP consultations, participants receive counselling, HIV/STI testing, and they provide information on demographics, sexual behaviour, STI history of the past year, and STI test results.

**Table S1.** Parameters for the three sexual activity groups among 250,000 MSM in the Netherlands.

| Parameter for activity group $i$ | Activity group | | | Source |
|---|---|---|---|---|
| | Low | Medium | High | |
| % in specific activity group | 60% | 35% | 5% | [a] |
| Rate of forming regular partnerships per year ($\alpha_{si}$) | 0.1 | 0.2 | 0.7 | [1] |
| Number of sex contacts per regular partner per day ($u_i$) | 0·2190 | 0·2320 | 0·3284 | [37] |
| % with regular partner ($q_i$) | 68% | 58% | 55% | [37] |
| Number of casual partners per day[b] ($\alpha_{ci}$) | 0·0202 | 0·0823 | 0·1-0.6 | [a] |

[a] Based on data from the Dutch national registration of Sexual Health Centre consultations (SOAP) [11] for MSM participating at the PrEP pilot in the Netherlands, from July 2019 (start of the PrEP pilot) until December 2022 (see Supplement for more details).

[b] The number of casual partners per day for the group with high level of sexual activity was included with a range in the uncertainty analysis.

**Table S2.** Characteristics of the three sexual activity groups: MSM with low, medium, or high sexual activity. Data of 8,500 MSM participating the PrEP pilot in the period July 2019 – December 2022, obtained from the Dutch national registration of Sexual Health Centre consultations (SOAP) [11].

| | **Sexual activity level** | | |
|---|---|---|---|
| | **Low** | **Medium** | **High** |
| **% of consultations** | 60% | 35% | 5% |
| **Number of consultations** | 36,854 | 22,072 | 2,859 |
| **Number of partners preceding 6 months, mean** | 3.64 | 14.81 | 78.18 |
| **Number of partners preceding 6 months, range** | 0-7 | 8-34 | 35-1,500 |

**Table S3.** Six-month transition probabilities between subgroups of MSM with different sexual activity levels. Data of 8,500 MSM participating the PrEP pilot in the period July 2019 – December 2022, obtained from the Dutch national registration of Sexual Health Centre consultations (SOAP) [11].

| | **Time *x* + 6 months** | | |
|---|---|---|---|
| **Time *x*** | **Low** | **Medium** | **High** |
| **Low** | 0.83 | 0.17 | 0.01 |
| **Medium** | 0.30 | 0.66 | 0.04 |
| **High** | 0.11 | 0.35 | 0.54 |

**Table S4.** Posterior distributions of uncertain model parameters obtained via the fitting process for the mpox outbreak among MSM in the Netherlands in 2022. Results shown are the medians and 95% credible intervals.

| **Model parameter** | **Median** | **95% range** | |
|---|---|---|---|
| Transmission probability per sex act, $\beta$ | 0.48 | 0.35 | 0.60 |
| Number of casual partners per day, for MSM with high sexual activity | 0.38 | 0.31 | 0.58 |
| Factor increasing transmission probability from symptomatic infectious case, compared to pre-symptomatic infectious, $w$ | 45% | 13% | 87% |
| Assortativeness in mixing with regular partners ($\varepsilon_r$) | 0.63 | 0.50 | 0.82 |
| Assortativeness in mixing with casual partners ($\varepsilon_c$) | 0.73 | 0.51 | 0.89 |
| Date of behavioural adaptations in June, $T_1$ | 19-6-2022 | 17-6-2022 | 24-6-2022 |
| Date of behavioural adaptations in July, $T_2$ | 8-7-2022 | 5-7-2022 | 14-7-2022 |
| Effective symptomatic infectious period (days), before $T_1$ | 16.0 | 10.0 | 20.4 |
| Effective symptomatic infectious period (days), between $T_1$ and $T_2$ | 4.9 | 2.8 | 7.9 |
| Effective symptomatic infectious period (days), after $T_2$ | 2.8 | 2.1 | 3.8 |
| % reduction in number of casual partners, between $T_1$ and $T_2$ | 34% | 1% | 49% |
| % reduction in number of casual partners, after $T_2$ | 32% | 1% | 49% |

**Table S5.** Scenarios for 2024-2026 with behavioural adaptations similar to those in 2022. Results show the cumulative number of mpox cases in the first four months after the introduction of 5 or 10 index cases in the population of 250,000 MSM in the Netherlands.

| Mpox vaccinations after 2023 | | Introduction of new mpox cases | | Number of mpox cases | |
|---|---|---|---|---|---|
| **Number of vaccinations** | **Timing of vaccination** | **Number of new introductions** | **Timing of new introductions** | **Median** | **Interquartile range** |
| 0 | - | 5 | 1 May 2025 | 759 | 492-1,050 |
| | | | 1 Nov 2025 | 863 | 569-1,177 |
| | | 10 | 1 May 2025 | 1,455 | 954-2,020 |
| | | | 1 Nov 2025 | 1,655 | 1,098-2,252 |
| 3,000 | Aug-Oct 2024 | 5 | 1 May 2025 | 502 | 307-722 |
| | | | 1 Nov 2025 | 652 | 418-922 |
| | | 10 | 1 May 2025 | 977 | 601-1,403 |
| | | | 1 Nov 2025 | 1,265 | 812-1,777 |
| | Feb-Apr 2025 | 5 | 1 May 2025 | 395 | 231-586 |
| | | | 1 Nov 2025 | 573 | 357-815 |
| | | 10 | 1 May 2025 | 769 | 453-1,150 |
| | | | 1 Nov 2025 | 1,113 | 697-1,577 |
| 30,000 | Aug-Oct 2024 | 5 | 1 May 2025 | 19 | 5-51 |
| | | | 1 Nov 2025 | 56 | 23-114 |
| | | 10 | 1 May 2025 | 39 | 11-101 |
| | | | 1 Nov 2025 | 111 | 47-229 |
| | Feb-Apr 2025 | 5 | 1 May 2025 | 6 | 0-25 |
| | | | 1 Nov 2025 | 22 | 6-55 |
| | | 10 | 1 May 2025 | 13 | 0-51 |
| | | | 1 Nov 2025 | 44 | 13-109 |

**Table S6.** Scenarios for 2024-2026 with (a) different behavioural adaptations than those in 2022 or (b) with a subclade with different characteristics than those of subclade IIb responsible for the 2022 outbreak. Results show the cumulative number of mpox cases in the first four months after the introduction of 5 index cases on 1 May or 1 November 2025, in the population of 250,000 MSM in the Netherlands. The scenarios were calculated without additional vaccinations after 2023.

| Scenario | | Timing of new mpox introductions | Number mpox cases | |
|---|---|---|---|---|
| | | | Median | Interquartile range |
| Behavioural adaptations in 2025, compared to 2022 | 14 days later | 1 May 2025 | 1,382 | 824-1,998 |
| | | 1 Nov 2025 | 1,616 | 976-2,273 |
| | 14 days earlier | 1 May 2025 | 375 | 263-500 |
| | | 1 Nov 2025 | 420 | 302-552 |
| | 25% lower level | 1 May 2025 | 1,000 | 648-1,422 |
| | | 1 Nov 2025 | 1,147 | 767-1,618 |
| | 25% higher level | 1 May 2025 | 632 | 423-859 |
| | | 1 Nov 2025 | 716 | 484-954 |
| | When number of cases exceeds 10 | 1 May 2025 | 62 | 48-85 |
| | | 1 Nov 2025 | 65 | 50-91 |
| | When number of cases exceeds 20 | 1 May 2025 | 118 | 95-173 |
| | | 1 Nov 2025 | 123 | 99-178 |
| Characteristics of new mpox clade (introduced in 2025), compared to subclade IIb responsible for the 2022 outbreak | 25% higher hospitalization rate | 1 May 2025 | 194 | 89-294 |
| | | 1 Nov 2025 | 219 | 102-329 |
| | 10% higher transmission probability | 1 May 2025 | 1,343 | 912-1,844 |
| | | 1 Nov 2025 | 1,543 | 1,070-2,084 |

**Figure S1.** Flow diagram of the model describing mpox transmission among men who have sex with men. Individuals in the model are divided into classes according to mpox status, vaccination status, and sexual activity level. Mpox status can be uninfected ($S_{ij}$), exposed not-infectious ($E_{ij}$), pre-symptomatic infectious ($I_{ij}$), symptomatic infectious ($Y_{ij}$), or recovered/removed ($R_{ij}$). The subscripts $i$ denote vaccination status: $i = 0$, unvaccinated; $i = 1$, partially vaccinated; $i = 2$, fully vaccinated. The subscripts $j = 1,2,3$ denote the groups with low, medium, or high sexual activity, respectively. In the model, we accounted for exit out of the population at a per capita rate $\mu$ that is not shown in the diagram. The parameters are defined in Table 1.

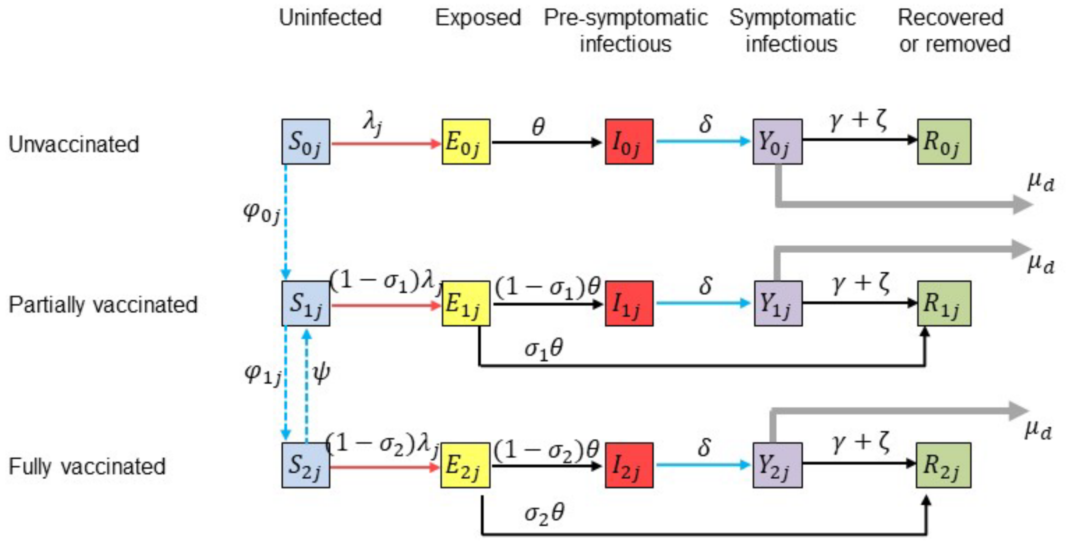

**Figure S2.** Data and model results for 2022-2023 in the Netherlands. Top panel: the number of mpox cases per day according to date of symptom onset. Black dots: data from the national surveillance system of notifiable infectious diseases of the Netherlands. Red and black lines: median and 95% credible intervals, calculated from the model. Bottom panel: the number of mpox vaccinations. Black dots: actual numbers of 1st and 2nd dose mpox vaccinations from data from the National Institute of Public Health and the Environment of the Netherlands. Red and blue line: the number of 1st and 2nd dose, respectively, of mpox vaccinations in the model.

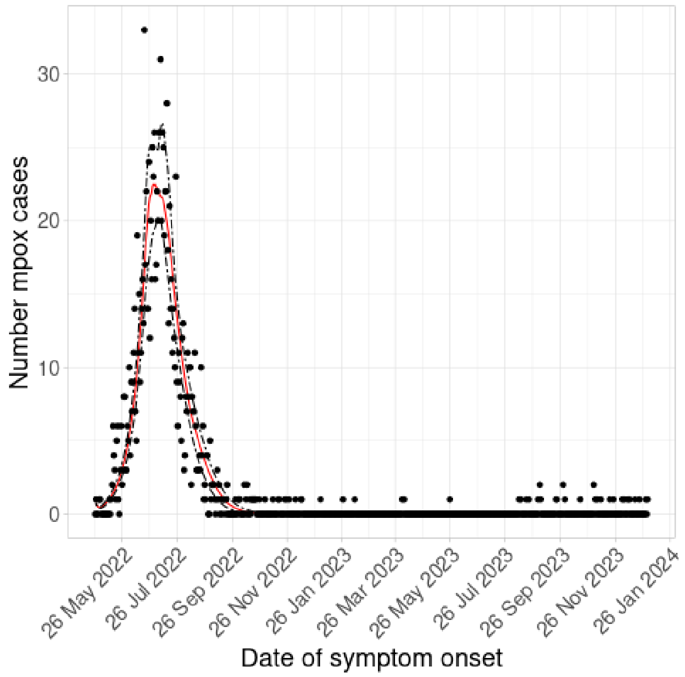


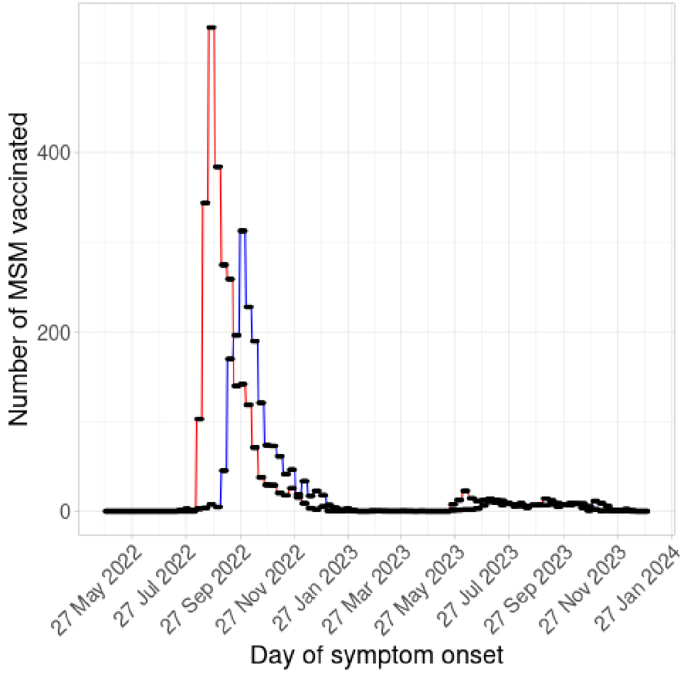

**Figure S3.** Mpox outbreaks after the introduction of 10 new mpox cases on 1 May (left panels; solid lines) or 1 November 2025 (right panels; dashed lines). The lines show the medians and the shaded areas show the 95% credible intervals for the scenarios with the respective colours. Black lines and grey shaded areas: without vaccinations after 2023. Top panels: red lines and shaded areas: with 3,000 vaccinations carried out in August-October 2024; dark blue lines and shaded areas: with 30,000 vaccinations carried out in August-October 2024. Bottom panels: pink lines and shaded areas: with 3,000 vaccinations carried out in February-April 2025; light blue lines and shaded areas: with 30,000 vaccinations carried out in February-April 2025. These scenarios are similar to those in Figure 2 in the main text, the only difference being the number of new mpox introductions: 5 cases in Figure 2, 10 cases in this figure.

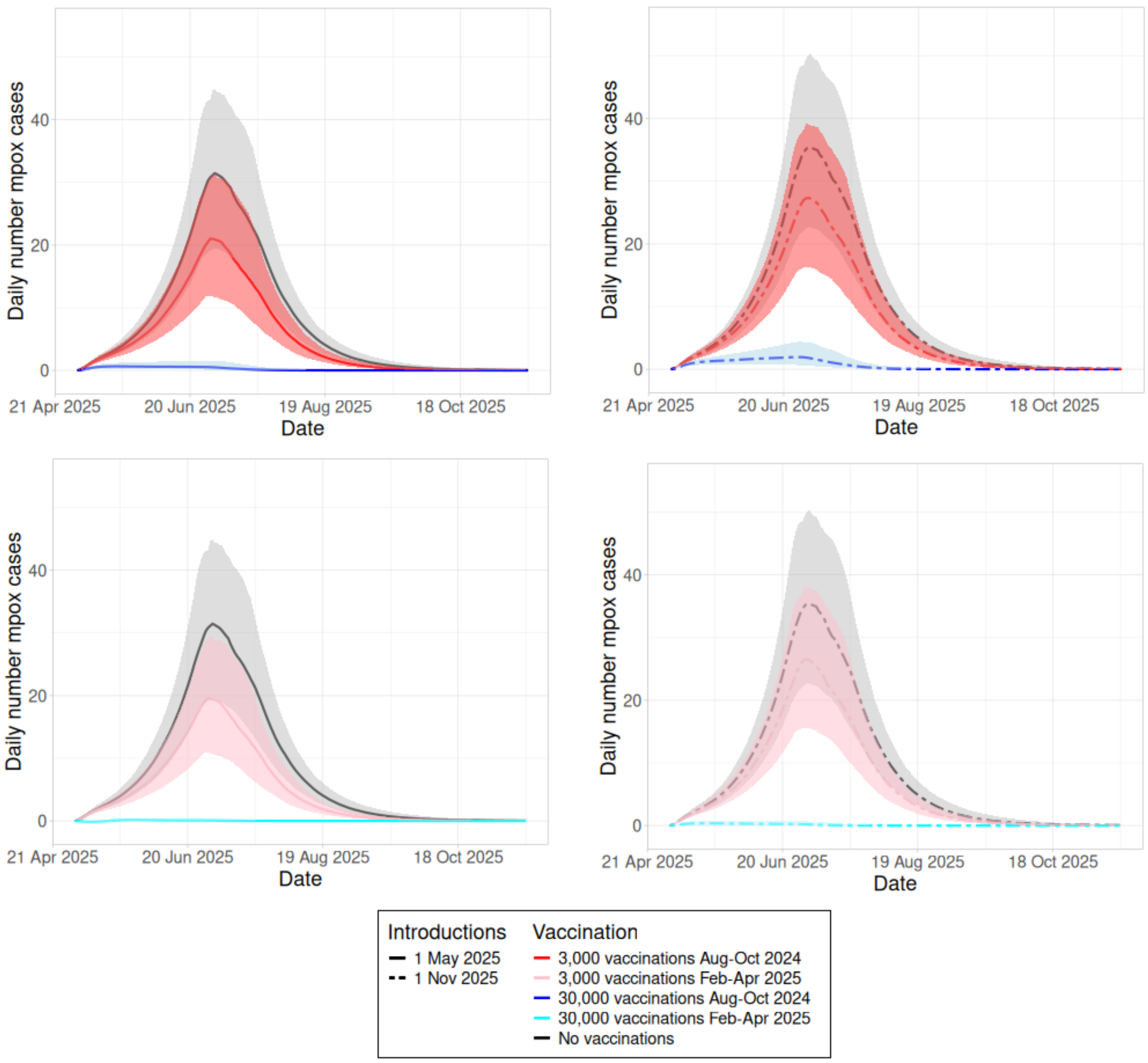

**Figure S4.** Mpox outbreaks after the introduction of 5 new (index) mpox cases on 1 May (left panel; solid lines) or 1 November 2025 (right panel; dashed lines) without vaccinations after 2023. The lines show the medians and the shaded areas show the 95% credible intervals for the scenarios with the respective colours. Black lines and grey shaded areas show the scenarios with behavioural adaptations as during the 2022 outbreak. Blue and read lines and shaded areas show the scenarios with behavioural adaptations starting when the number of symptomatic infectious cases exceeded 10 cases (red) or 20 cases (blue).

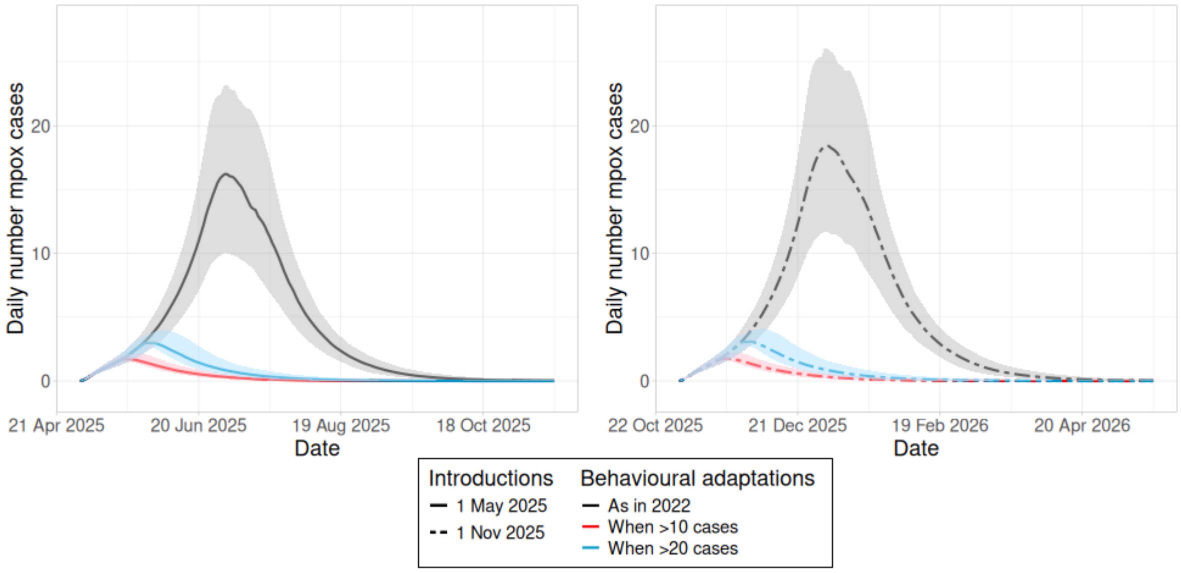

**Figure S5.** Mpox outbreaks after the introduction of 10 (top panels) or 20 (bottom panels) new mpox cases on 1 May (solid lines in left panels) or 1 November 2025 (dashed lines in right panels), without additional vaccinations after 2023. The lines show the medians and the shaded areas show the 95% credible intervals for the scenarios with the respective colours. The new cases were infected with a subclade having: the same properties as subclade IIb, responsible for the 2022 outbreaks (black lines and grey shaded areas); or with higher infectivity (red lines and pink shaded areas); or with higher hospitalization rate (blue lines and shaded areas). .

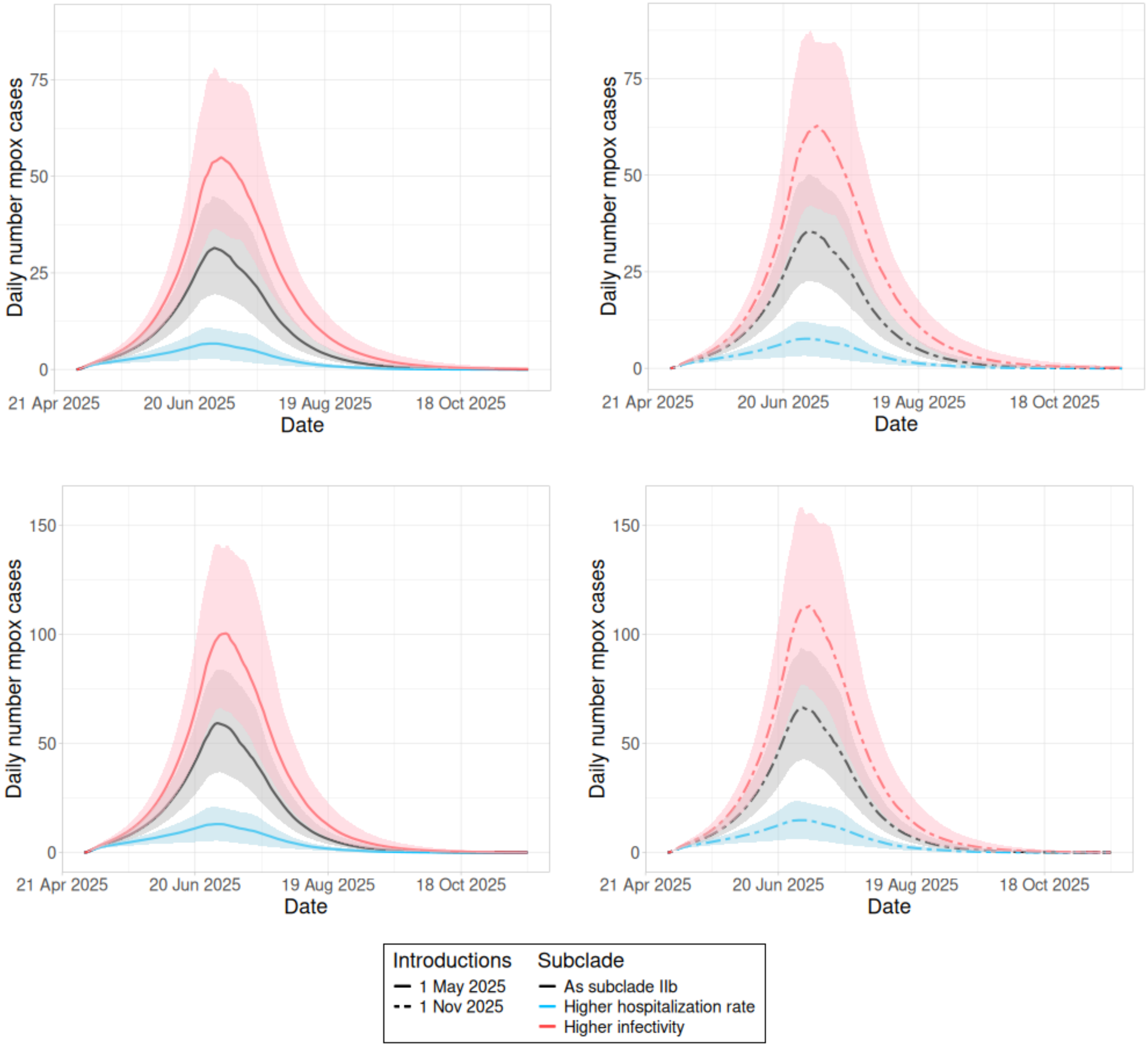

19. National Institute of Public Health and the Environment (RIVM). Epidemiological situation of mpox in the Netherlands, update 3 April 2023. Bilthoven, 2023. Available from:

20. National Institute of Public Health and the Environment (RIVM) STI Department. Sexually transmitted infections in the Netherlands in 2020. Report number 2021-0052. Bilthoven, 2021. Available from:

https://rivm.openrepository.com/bitstream/handle/10029/625007/2021-0052.pdf. Accessed 18 November 2024.

28. Chow EPF, Samra RS, Bradshaw CS, Chen MY, Williamson DA, Towns JM, *et al*. Mpox knowledge, vaccination and intention to reduce sexual risk practices among men who have sex with men and transgender

people in response to the 2022 mpox outbreak: a cross-sectional study in Victoria, Australia. *Sex Health* 2023;**20**:390-402.
